# Investigation of convergent and divergent genetic influences underlying schizophrenia and alcohol use disorder

**DOI:** 10.1101/2020.10.27.20220186

**Authors:** Emma C. Johnson, Manav Kapoor, Alexander S. Hatoum, Hang Zhou, Renato Polimanti, Frank R. Wendt, Raymond K. Walters, Dongbing Lai, Rachel L. Kember, Sarah Hartz, Jacquelyn L. Meyers, Roseann E. Peterson, Stephan Ripke, Tim B. Bigdeli, Ayman H. Fanous, Carlos N. Pato, Michele T. Pato, Alison M. Goate, Henry R. Kranzler, Michael C. O’Donovan, James T.R. Walters, Joel Gelernter, Howard J. Edenberg, Arpana Agrawal

## Abstract

**Background:** Alcohol use disorder (AUD) and schizophrenia (SCZ) frequently co-occur, and recent genome-wide association studies (GWAS) have identified significant genetic correlations between them.

**Methods:** We used the largest published GWAS for AUD (total cases = 77,822) and SCZ (total cases = 46,827) to systematically identify genetic variants that influence both disorders (in either the same or opposite direction of effect) as well as disorder-specific loci.

**Results:** We identified 55 independent genome-wide significant SNPs with the same direction of effect on AUD and SCZ, 8 with robust effects in opposite directions, and 98 with disorder-specific effects. We also found evidence for 12 genes whose pleiotropic associations with AUD and SCZ are consistent with mediation via gene expression in the prefrontal cortex. The genetic covariance between AUD and SCZ was concentrated in genomic regions functional in brain tissues (p = 0.001).

**Conclusions:** Our findings provide further evidence that SCZ shares meaningful genetic overlap with AUD, and suggest that genetic variants with an effect on both disorders are expressed in brain tissues.

## Introduction

Schizophrenia (SCZ) and alcohol use disorder (AUD) are serious psychiatric disorders^1^. AUD is more common in individuals with SCZ (prevalence of 20-30%^2,3^, compared to ∼6% in the general population^4^) and a diagnosis of both is associated with greater psychiatric comorbidity^5^, more clinical complications^6^, and a lower likelihood of sustained medication adherence^7^ than either disorder alone. Both SCZ and AUD are moderately to highly heritable (twin-*h*^*2*^ for SCZ = 81%^8^, AUD = 49%^9^), and genome-wide association studies (GWAS) have consistently found positive genetic correlations of AUD with SCZ (e.g., *r*_*g*_ = 0.34, p = 3.7e-21)^10^. Further, polygenic risk scores (PRS) for AUD are significantly associated with SCZ risk^10^ and vice versa^11^. In contrast, the genetic correlation between SCZ and measures of typical alcohol consumption is weak (e.g., drinks/week: *r*_*g*_ = 0.01, p = 0.67^12^), suggesting that SCZ might share substantial genetic liability only with the psychopathological aspects of disordered drinking^13^ and not alcohol consumption *per se*.

Despite the substantial genetic correlation between AUD and SCZ, there is little known regarding the underlying pleiotropic mechanisms in terms of the specific risk alleles, genes, and molecular pathways involved. Although recent studies have begun to elucidate the contributions of pleiotropic loci to shared genetic variance amongst disorders and complex traits^14,15^, these efforts have not included AUD, one of the most common psychiatric disorders. In addition to loci with a similar direction of effect on both disorders, modern cross-disorder GWAS methods can also identify divergent variants, i.e., those that are pleiotropic for two disorders but whose effect alleles operate in opposite directions, conferring risk for one disorder and protective effects for the other (e.g., potassium ion response genes that distinguish SCZ from bipolar disorder^16^). The identification of such variants is fundamental to identifying the pathways that contribute to diagnostic boundaries.

The current study outlines the nature of the shared genetic underpinnings of AUD and SCZ by conducting cross-disorder analyses of large genome-wide datasets of both European- and African-ancestry individuals (see **Figure 1** for overview). We conducted ancestry-specific cross-disorder meta-analyses to systematically identify pleiotropic loci with significant convergent and divergent effects on both SCZ and AUD, and loci specific to each disorder. We also linked pleiotropic variants to gene expression data from the frontal cortex in an effort to prioritize the genes that are more likely to be causal. Because the extent to which the correlation between AUD and SCZ is attributable to salient functional categories remains unknown, we partitioned the genetic covariance between AUD and SCZ into relevant annotations.

**Figure 1.**
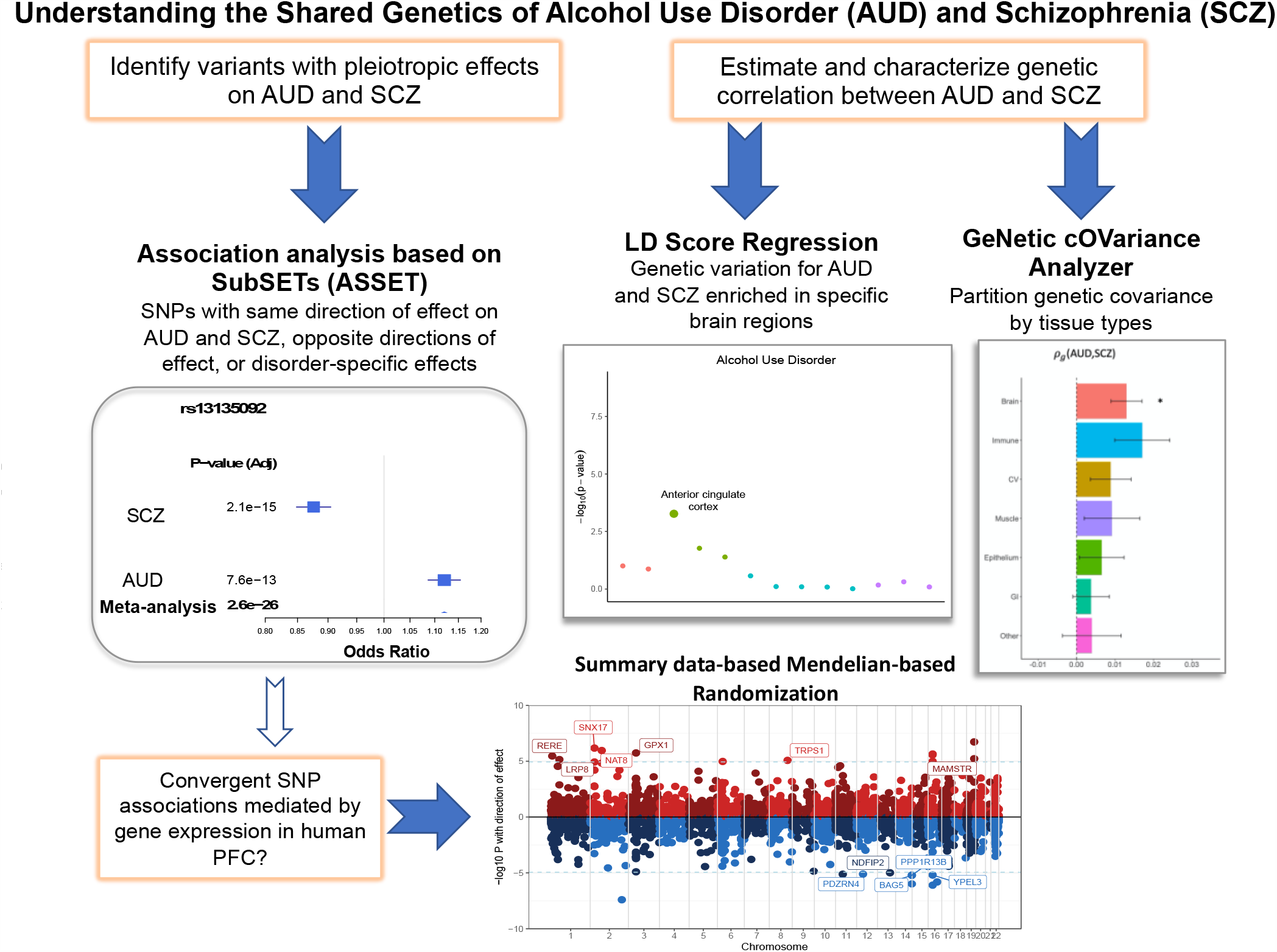
Conceptual overview of cross-disorder analysis of alcohol use disorder and schizophrenia.

## Methods and Materials

### Samples

#### Alcohol Use Disorder

For the European-ancestry subset, we utilized GWAS data from the largest available AUD meta-analysis^17^ (N = 313,959; N_cases_ = 57,564). This study meta-analyzed AUD GWAS data from the Million Veteran Program (MVP), where case status was derived from International Classification of Diseases (ICD) codes of alcohol-related diagnoses in electronic health records (EHR) data, and the Psychiatric Genomics Consortium (PGC) alcohol dependence GWAS^18^ with cases based on DSM-IV diagnoses.

We meta-analyzed two published GWAS ourselves to create the African-ancestry subset: the MVP Phase 1 GWAS of AUD^10^ (N = 56,648; N_cases_ = 17,267), and the PGC GWAS^18^ (N = 5,799; N_cases_ = 2,991). We used METAL^19^ (which combines genome-wide summary statistics across multiple samples) to generate the African-ancestry summary statistics by meta-analyzing the GWAS data from the MVP Phase 1 AUD and PGC alcohol dependence GWAS using an inverse variance-weighted fixed-effects model, excluding SNPs with INFO score < 0.8 and/or minor allele frequency < 0.01 within each sample.

#### Schizophrenia

For the European-ancestry sample, we used the PGC Phase 2 + CLOZUK (a sample of individuals with schizophrenia who were treated with clozapine) SCZ GWAS meta-analysis^20^ (total N = 105,318; N_cases_ = 40,675). We used the summary statistics from the Genomic Psychiatry Cohort (GPC) SCZ GWAS^21^ (N = 10,070; N_cases_ = 6,152) for the African-ancestry cross-disorder analysis.

### Analysis

#### Cross-disorder association analysis

We used “Association analysis based on SubSETs” (ASSET)^22^ to combine the genome-wide association data for AUD and SCZ (separately by ancestry), using the two-tailed meta-analysis approach to obtain a single cross-disorder association statistic, correcting for sample overlap. Unlike traditional meta-analysis approaches, ASSET takes into account SNPs with significant effects on multiple disorders even if the effects on the traits are in opposite directions. Default parameters were applied using the “h.traits” function, and we used LD Score Regression^23,24^ (LDSC) to obtain a rough estimate of sample overlap, which we accounted for in the additional covariance term. We then separated the ASSET results into four subsets: a “convergent” subset (effect allele with the same direction of effect for both disorders), a “divergent” subset (effect allele with opposite directions of effect on the disorders), a subset of SNPs with AUD-specific effects, and a subset of SNPs with effects only on SCZ (**Supplemental Figure 1**).

In the European-ancestry sample, we then uploaded the subset results (i.e., convergent, divergent, AUD-only, and SCZ-only SNPs from ASSET output) to FUMA v1.3.6a^25^ for annotation and identification of genome-wide significant risk loci and independent lead SNPs. We further subset these convergent and divergent loci to exclude top lead SNPs with p > 0.05 in either individual disorder GWAS, to create a more conservative set of cross-disorder variants with at least nominal significance in both disorders (although the full subsets were used for gene-set and pathway analyses).

In the African-ancestry samples, which were smaller and accordingly lacked power for more extensive analyses, we focused on the overall set of pleiotropic cross-disorder variants, rather than parsing the pleiotropic variants into subsets with convergent and divergent effects. In addition, as much of the data required for follow-up analyses described below were restricted to European-ancestry individuals, these follow-up analyses were not conducted on the African-ancestry cross-disorder summary statistics.

#### Gene, gene-set, and pathway analyses

In the European-ancestry samples, gene-based analyses in MAGMA (v1.08)^26^ were conducted on the subset results (i.e., convergent and divergent SNPs from ASSET output) via the FUMA^25^ platform. These analyses included gene-set analyses using curated gene sets and GO terms from MsigDB^27^ (an online collection of annotated gene-sets), gene-property analyses based on tissue expression data from GTEx^28^ v8 (a repository of expression data by brain region from autopsies of 960 donors), and data visualization (more details in **Supplemental Materials**).

#### SMR eQTL analyses and differential gene expression analyses

To examine whether the effects of pleiotropic variants with convergent effects on AUD and SCZ may be mediated by gene expression patterns, we conducted a summary data-based Mendelian randomization^29^ (SMR) analysis on a set of expression quantitative trait loci (eQTL) data in the prefrontal cortex (meta-analyzed to combine data from ROSMAP^30^, PsychENCODE^31^, and COGA-INIA datasets^32^; total N = 1,986). SMR is a Mendelian randomization-based analysis that integrates GWAS summary statistics with eQTL data to test whether the effect size of a SNP on the phenotype of interest is mediated by gene expression. SMR does not require raw eQTL data to build the weights. We excluded variants with pleiotropic effects significantly different from what would be expected under a causal model using the HEIDI-outlier method^33^ (excluding SNPs with HEIDI-outlier p < 0.05).

We also examined whether genes mapped to pleiotropic loci by MAGMA (with p < 0.05) were significantly enriched for genes showing differential expression (p < 0.05) in the prefrontal cortex using two comparisons in independent samples: we compared gene expression in 65 individuals with alcohol dependence and 73 healthy controls^32^, and 258 individuals with SCZ and 279 controls^34^ (details in Supplemental Note) using Fisher’s exact test^35^.

#### Genetic correlations and partitioned covariance

We used LDSC^23,24^ to estimate the genetic correlations (r_g_) between AUD, SCZ, and two negative control traits (height and chronic ischemic heart disease). We compared the genetic correlations between AUD and SCZ, and DPW and SCZ using a block jackknife method implemented through LDSC to test the null hypothesis that r_g_(AUD, SCZ) minus r_g_(DPW, SCZ) = 0.

We also used LDSC applied to Specifically Expressed Genes (LDSC-SEG^36^) to estimate the enrichment of AUD and SCZ across 13 specific brain regions (annotations defined using GTEx^37^ gene expression data).

We used GeNetic cOVariance Analyzer (GNOVA)^38^ to partition the genetic covariance (*ρ*_*g*_) between AUD and SCZ into salient annotation categories. These included 1) functional vs. non-functional areas of the genome (GenoCanyon^39^ annotations, defined by integrating genomic conservation measures and biochemical annotation data to generate a functional potential score for each genetic variant), 2) tissue- and regional-specific functionality (GenoSkyline^40,41^ annotations, which are tissue-specific functional regions defined by integrating high-throughput epigenetic annotations, 3) GTEx v6^37^ brain region annotations from LDSC cell-type specific analyses), and 4) minor allele frequency quartiles. We excluded the MHC region (chr6:26000885 - chr6:33999991) from both the LDSC-SEG and GNOVA analyses due to the long-range and complex LD in this region.

In sensitivity analyses conducted on individuals of European ancestry in the UK Biobank (from the Neale lab GWAS: https://www.nealelab.is/uk-biobank), we calculated the genetic correlation and partitioned genetic covariance between AUD, SCZ, and two negative control traits: height (N = 360,388) and chronic ischemic heart disease (CHD; N = 361,194; N_cases_ = 12,769).

## Results

### Identifying pleiotropic variants, genes, and pathways in individuals of European ancestry

The cross-disorder analysis of AUD and SCZ in European-ancestry individuals identified numerous significant pleiotropic loci: after genome-wide clumping via FUMA^25^, there were 55 independent risk loci (with 60 lead SNPs) with convergent effects and 44 risk loci (56 lead SNPs) with divergent effects (i.e., effect allele with opposite effects on AUD and SCZ; **Supplemental Figure 1, Supplemental Tables 1-3)**. We also identified disorder-specific loci through ASSET: 90 with SCZ-only effects, and 8 with AUD-only effects (**Supplemental Tables 4-5**). MAGMA gene-based analyses identified 119 significant genes from the convergent subset and 105 genes from the divergent subset (**Supplemental Figure 2, Supplemental Tables 6-7**).

**Figure 2.**
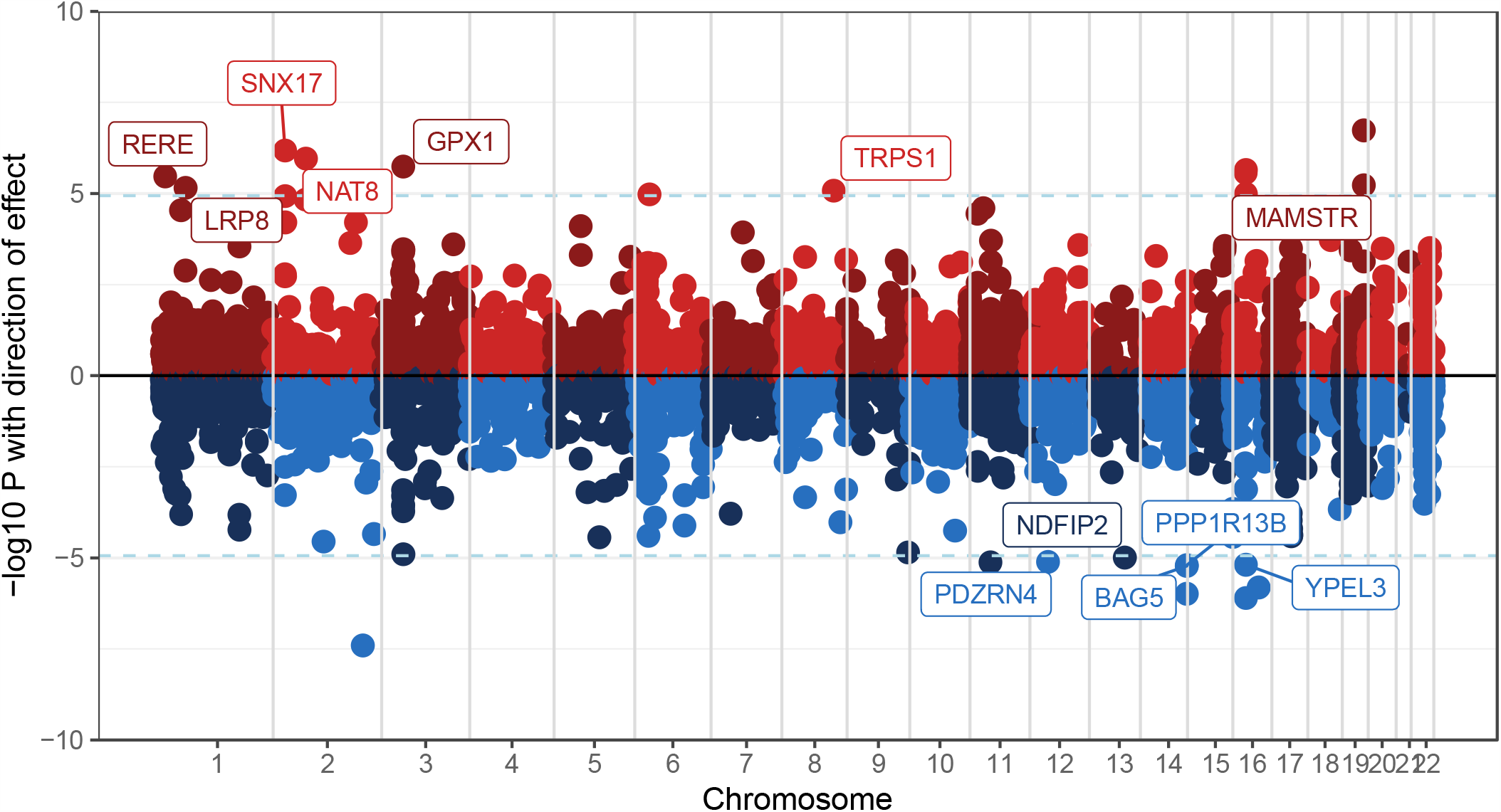
Genes in the convergent subset whose association with AUD and SCZ may be mediated by gene expression in the prefrontal cortex, analyzed using Summary data-based Mendelian Randomization (SMR). Genes in red show up-regulated gene expression, and genes in blue show down-regulated gene expression. The 12 labeled genes are significant after Bonferroni corrections and were not excluded as pleiotropic outliers (HEIDI-outlier method p > 0.05)

As ASSET^22^ searches for and determines the most likely subset for each SNP (i.e., classifying SNPs as having an effect only on AUD, only on SCZ, or on both disorders), some of the pleiotropic SNPs identified by ASSET were only significant in one of the single-disorder GWAS. For a more conservative description of pleiotropic loci of divergent effect, we further considered only the 8 loci where the top lead SNPs had opposite effects but p < 0.05 for both AUD and SCZ (**Supplemental Table 3**; the convergent lead SNPs already had p < 0.05 for both disorders). In the convergent subset, the strongest association was on chromosome 11 (lead SNP rs6589386, cross-disorder p = 5.7e-18; AUD p = 7.1e-12, SCZ p = 1.6e-8). The strongest divergent signal was on chromosome 4 (lead SNP rs13135092, cross-disorder p = 2.9e-31; AUD p = 4.9e-18, SCZ p = 7.9e-16), located in an intron of *SLC39A8*. Importantly, although rs13135092 is pleiotropic for both AUD and SCZ, the same effect allele (A) *increases risk for* AUD and *decreases risk* (i.e., is protective) for SCZ.

MAGMA competitive gene-set analyses identified 2 significant GO terms in the convergent subset of variants, one related to DNA binding (GO: 0043565, p = 1.3e-6) and one related to neuronal differentiation (GO: 0045664, p = 1.9e-6; **Supplemental Table 8**). There were no significant gene-sets identified for the divergent subset).

Both the convergent and divergent subsets of variants showed enrichment in all 13 brain tissues in MAGMA gene-property analyses, and the divergent subset also showed enrichment in pituitary tissues. (**Supplemental Figure 3**).

**Figure 3.**
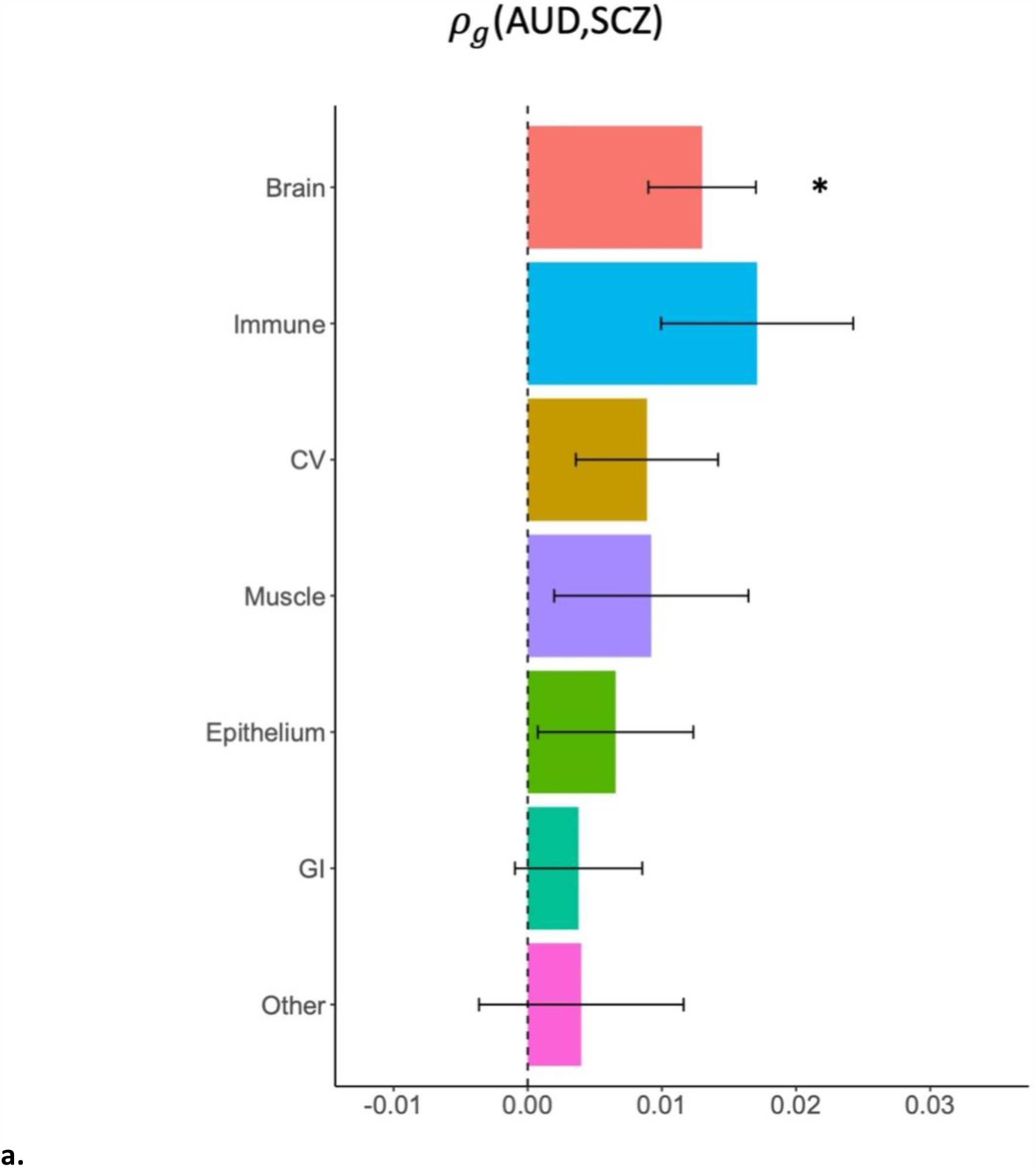
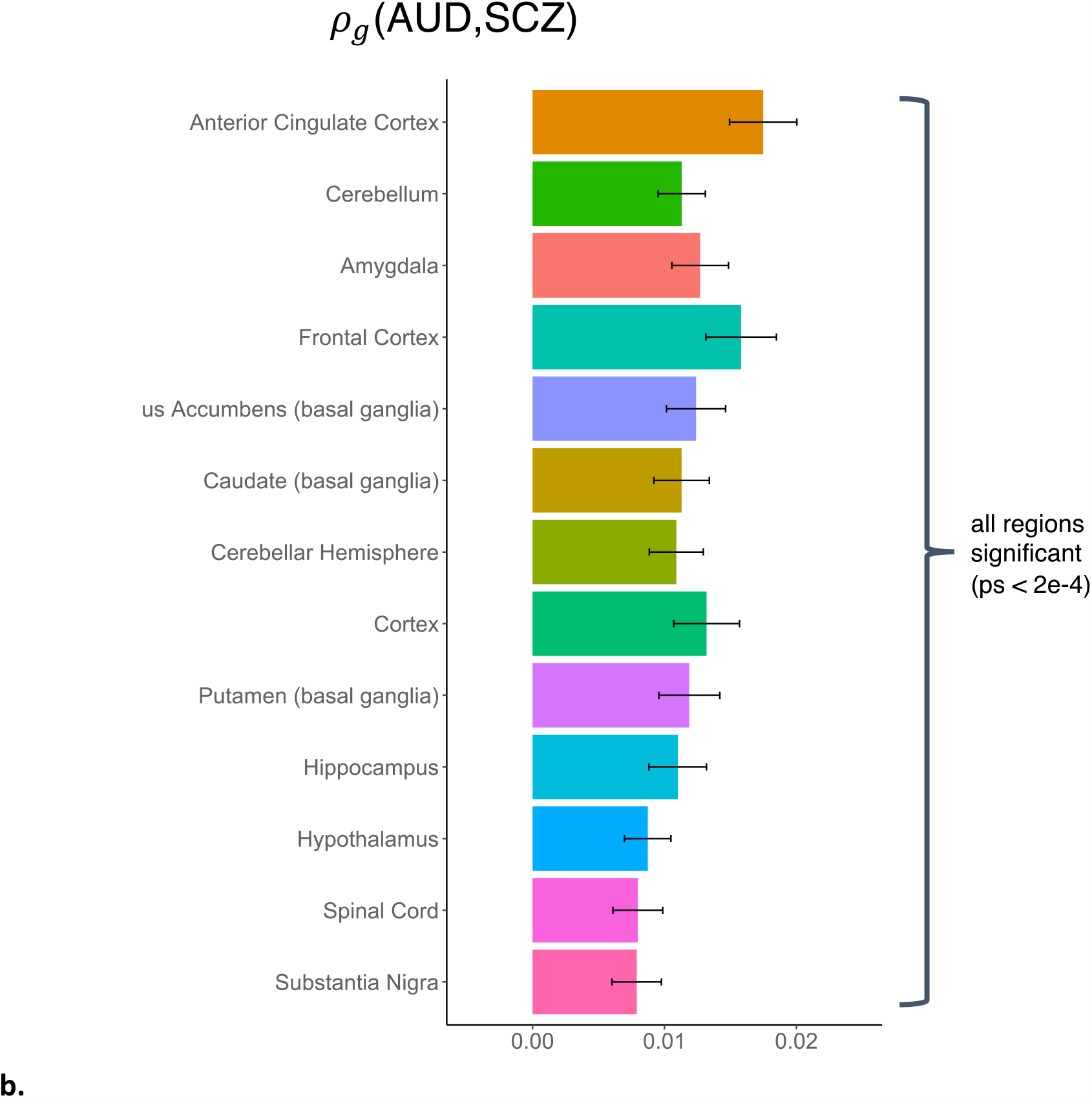
Stratified genetic covariance between AUD and SCZ. **a:** stratified by broad tissue type. Tissue annotations were defined using the GenoSkyline-Plus annotations. Significant tissues are starred. CV = cardiovascular; GI = gastrointestinal. **b:** stratified by 13 brain regions, defined using GTEx v6 gene expression data.

### Cross-disorder variants in African ancestry individuals

In the African-ancestry cross-disorder analysis, there was limited power to identify pleiotropic loci, with results appearing to be primarily driven by the larger AUD GWAS (e.g., the one genome-wide significant SNP that ASSET identified as being pleiotropic had p > 0.05 for SCZ).

### Integration of eQTL data and differential gene expression in EUR-ancestry samples

Of the 22 genes that survived Bonferroni correction (for the number of genes tested; p_SMR_ < 1.16e-5), SMR analyses identified 12 convergent genes whose cross-disorder associations with AUD and SCZ were consistent with mediation via gene expression in the prefrontal cortex (PFC; the remaining 10 genes with p_SMR_ < 1.16e-5 had p_HEIDI_ < 0.05, indicating pleiotropy outlier status; **Figure 2**; significant results in **Supplemental Table 10;** full results in **Supplemental Table 11**).

Neither the genes with variants of convergent nor divergent effect were enriched in specific differential gene expression analyses of postmortem PFC tissue of individuals with SCZ (N_cases_ = 258) vs. controls or of AUD (N_cases_ = 65) vs. controls.

### Genetic covariance and correlation

There were significant positive genetic correlations (r_g_) between AUD and SCZ (r_g_ = 0.392, p = 1.2e-42). In sensitivity tests, the negative control measure of chronic ischemic heart disease was not significantly correlated with AUD but height showed a small negative correlation (r_g_ = -0.093, SE = 0.021, p = 7.54e-6). SCZ showed a nominal genetic correlation with heart disease (r_g_ = -0.063, SE = 0.029, p = 0.032), but none with height.

LDSC-SEG analyses that partitioned heritability enrichment by tissue-specific gene expression revealed significant enrichment for SCZ in three of 13 brain regions: the cortex, frontal cortex, and anterior cingulate cortex (FDR q-values < 2e-5), and for AUD in the anterior cingulate cortex (FDR q-value = 0.007; **Supplemental Figure 4**).

The genetic covariance (*ρ*_*g*_) of AUD and SCZ was significantly attributable both to functional regions of the genome (p = 1.0e-8), including those specifically functional in brain tissues (p = 0.001; see **Figure 3a, Supplemental Table 12**) and non-functional regions (p = 1.5e-19; **Supplemental Table 13**), as well as all minor allele frequency quartiles except the lowest frequency quartile (p = 2.9e-5 - 3.9e-9; **Supplemental Table 14**). While the point estimate of genetic covariance was highest in genes functional in immune tissues (*ρ*_*g*_ = 0.017), this estimate had a relatively large standard error (0.007) and did not reach significance after multiple testing corrections. There were no significant findings when partitioning the genetic covariance between AUD and height or SCZ and heart disease into tissue-specific categories with GNOVA.

Because the genetic covariance of AUD and SCZ was significantly concentrated in brain tissues, we partitioned it further into specific brain regions, using the 13 GTEx brain region annotations provided by LDSC-SEG^36^. We found significant concentrations of genetic covariance in all 13 brain regions tested, with the greatest concentrations in the anterior cingulate cortex (p = 7.6e-12), frontal cortex (p = 3.3e-9), cortex (p = 1.2e-7), and amygdala (p = 3.1e-9; **Supplemental Table 15, Figure 3b**).

## Discussion

The prevalence of AUD is elevated in those with SCZ, relative to the general population. Recent large GWAS of AUD document robust genetic correlations between AUD and SCZ^17^. Utilizing a subset-based meta-analytic approach, our cross-disorder analysis of AUD and SCZ identified loci with either the same or opposite direction of effects on AUD and SCZ; 55 convergent loci and 8 divergent loci had lead SNPs that were p < 0.05 in both individual disorders. The genetic covariance between AUD and SCZ was concentrated in genes functional in the brain, and eQTL analyses of the convergent subset of variants (i.e., those with the same direction of effect on both disorders) identified 12 genes whose association with AUD and SCZ may be mediated via gene expression in brain tissues.

AUD and SCZ were robustly genetically correlated. In contrast, prior studies have found that SCZ is uncorrelated with measures of typical alcohol consumption (e.g., with drinks/week, r_g_ = 0.01, p = 0.67^12^), especially when alcohol frequency of use alone is considered^42^. Genetic correlations with indices that encompass heavy episodic drinking, such as the consumption sub-scale of the Alcohol Use Disorders Identification Test (AUDIT-C), are also lower (r_g_ = -0.0003 - 0.04)^10,13^ than those noted for AUD. A recent study has implicated that indices of socioeconomic status may influence genetic correlations between measures of substance use and psychopathology^43^; however, even after co-varying for SES, the genetic correlation between alcohol frequency and SCZ remained non-significant. The genetic correlation between alcohol quantity and SCZ remained significant and relatively unchanged (r_g_ = 0.14-0.16), but markedly lower than the AUD-SCZ genetic correlation. This suggests that SCZ shares genetic variation primarily with the psychopathological aspects of problem drinking and AUD, even after accounting for socioeconomic measures.

The top *divergent* SNP, rs13135092, in an intron of *SLC39A*, showed strong associations with both AUD (p = 4.9e-18) and SCZ (p = 7.9e-16), with the A allele exerting a risk-increasing effect on AUD and a protective effect on SCZ. Comparison of summary statistics for over 4,756 traits^44^ indicated that the A allele was also associated with greater alcohol consumption, greater risk-taking, higher waist-hip ratio, lower bioelectrical impedance (i.e., greater adiposity) and higher systolic blood pressure, consistent with the direction of genetic association between these measures and AUD. However, the A allele was also associated with increasing cognitive performance, higher intelligence and with higher educational attainment – while the direction of these effects is consistent with the “protective” effect of the A allele on risk for SCZ, it contradicts prior research showing an inverse genetic correlation between educational attainment and AUD. Given the convergent direction of associations with AUD and alcohol consumption, as well as aspects of risk-taking and cardio-metabolic traits, but divergent direction of associations with SCZ as well as cognition, we speculate that the A allele of this SNP may be related to milder AUD, typified by positive reward-related drinking and impulsivity that is effectively regulated by enhanced cognitive functioning.

Our top convergent SNP, rs6589386 is intergenic and an eQTL for *DRD2* in cerebellar hemisphere tissue; this gene has been implicated for both AUD and SCZ across recent GWAS. In addition to AUD and SCZ, this variant has been implicated in GWAS of neuroticism, subjective well-being, alcohol consumption, and cigarette smoking.

Linking cross-disorder associations to eQTLs in the prefrontal cortex (PFC) suggested that the effects of pleiotropic variants on AUD and SCZ may be mediated by expression levels of several genes (**Figure 2**). Several of the identified genes were previously implicated in GWAS of metabolic traits (including *NAT8, TRPS1* (up-regulated), *BAG5, PPP1R13B* (down-regulated)), immunological traits (e.g., *RERE, TRPS1*, both up-regulated) and psychiatric phenotypes (e.g., up-regulated: *LRP8*, down-regulated: *PPP1R13B*). Although these findings may not be limited to gene expression in the PFC, the limited data currently available do not permit examination of whether these findings extend to other brain regions.

While there are limited samples available with measured gene expression in post mortem brain tissue, particularly for substance use disorders, we used two independent datasets (alcohol dependent and control samples from New South Wales Tissue Resource Centre at the University of Sydney, schizophrenia and control samples from the CommonMind consortium) to link pleiotropic variants to differential gene expression. We found no evidence for enrichment of genes pleiotropically associated with both AUD and SCZ when considering genes differentially expressed in the brains of individuals with AUD or SCZ (compared to respective controls). However, our lack of findings could be due to limited statistical power in the RNAseq whole-genome transcriptome datasets (N = 138 - 537) relative to the GWAS datasets (Ns > 100,000).

There are several limitations of the current study. First, nearly all gene expression reference datasets and other bioinformatic resources have been trained on European-ancestry samples. In addition, our African-ancestry samples were under-powered relative to the European-ancestry samples, and the SCZ sample more so than the AUD sample. Thus, the African-ancestry results were limited to the ASSET cross-disorder analysis, and appeared to be driven almost entirely by the larger AUD GWAS. Our motivation for conducting this cross-disorder analysis is because most cross-disorder GWAS rely on European-ancestry samples, generating inequity in downstream analyses of non-European populations. For instance, summary statistics from this cross-disorder GWAS could be used to generate polygenic risk scores in independent cohorts of African-American participants. Nonetheless, vast increases in African ancestry sample sizes are needed for studies of serious psychiatric conditions. Second, given the available data, we were limited to studying common genetic variation (minor allele frequencies > 1%); thus, there may be rare variants of importance underlying the comorbidity between AUD and SCZ that are outside the scope of the current study. Third, in our sensitivity analysis, we found a significant negative genetic correlation between AUD and height (r_g_ = - 0.093, SE = 0.021, p = 7.54e-6), which is likely driven in part by the very large sample sizes and polygenic nature of both traits, though there is some evidence for this relationship in the literature. One previous study found a negative genetic correlation between alcohol consumption and childhood height in males^45^ (r_g_ = -0.23, p = 0.002) and another study found a negative correlation between problematic alcohol use and comparative height at age 10 (r_g_ = - 0.09, p = 7.3e-5), though this was not significant when a Bonferroni correction was applied for the 715 traits tested^17^. Finally, while most of the AUD samples were screened for SCZ, some individuals among the SCZ samples very likely also had AUD. Unfortunately, there are no data currently available on which SCZ samples were screened for substance use disorders, to permit an estimation of such a bias.

Cross-disorder GWAS provide insights into genetic sources of comorbidity. While prior cross-disorder studies have provided foundational results for common psychiatric disorders, ours is amongst the first studies to focus on identifying pleiotropic loci for a substance use disorder, AUD, and SCZ. To understand the shared biology between substance use disorders and other psychiatric disorders, future efforts must include large numbers of individuals with AUD or problematic alcohol use rather than simple measures of consumption.

## Data Availability

Data from the PGC are available for download here: https://www.med.unc.edu/pgc/data-index/
Data from MVP are available for download from dbGAP.
Data from the GPC are available by request to the Genomic Psychiatry Cohort.
Data from Pardiñas et al. are available for download here: https://walters.psycm.cf.ac.uk

## ACKNOWLEDGEMENTS

The Psychiatric Genomics Consortium’s Substance Use Disorders (PGC-SUD) working group gratefully acknowledges prior support from NIAAA and thanks all our contributing investigators and study participants who make this research possible. We acknowledge collaborations with the Million Veteran Program, Psychiatric Genomics Consortium Substance Use Disorders Working Group, and Genomic Psychiatry Cohort Investigators. A list of Genomic Psychiatry Cohort investigators and the MVP Core Acknowledgement are provided in the Supplemental Materials. We are grateful to Luke Evans for helpful discussion of the manuscript.

