## Supplemental Note for "Investigation of convergent and divergent genetic influences underlying schizophrenia and alcohol use disorder"

**Methods**

***Meta-analysis of PGC Alcdep and MVP AUD African-ancestries samples***

We used METAL^1^ to meta-analyze the summary statistics from the MVP1 AUD^2^ and PGC alcohol dependence^3^ (Alcdep) African-ancestries GWAS using an inverse variance-weighted fixed-effects model, excluding SNPs with INFO score < 0.8 and/or minor allele frequency (MAF) < 0.01 within either sample.

***Association analysis based on SubSETs (ASSET)***

ASSET is a generalized version of fixed-effects meta-analysis; it allows a subset of the input GWAS to have no effect on a given SNP, and explores all possible subsets of “non-null” GWAS inputs to identify the strongest association signal in positive and/or negative directions. The final test-statistic thus accounts for both positive and negative (or null) directions of association at each SNP in the input file, combining the p-values from both positive and negative associations using Fisher’s method^4^ to create an overall Z-score and p-value. Thus, ASSET also permits the addition of a covariance term for the adjustment of overlapping samples. We selected ASSET for its ability to identify not only convergent pleiotropic variants (i.e. SNPs with the same direction of effect on each disorder) but also divergent pleiotropic SNPs, its conservative effect estimates and minimal inflation, and previous use in the literature^5^.

As ASSET searches for and determines the most likely subset for each SNP (i.e., classifying SNPs as having an effect only on AUD, only on SCZ, or having an effect on both disorders), the ASSET-identified pleiotropic SNPs were not necessarily significant in both single-disorder GWAS. For a more conservative description of pleiotropic loci, we further considered only the lead SNPs that had p < 0.05 in both disorders; after excluding SNPs with p $\geq$ 0.05 in either disorder, there were 8 divergent lead SNPs and 55 convergent lead SNPs (the lead SNPs in the genome-wide loci for the convergent subset already had p < 0.05 in both disorders; **Supplemental Table 3**).

***MAGMA analyses conducted through FUMA***

We used the FUMA^6^ v1.3.6a platform to conduct gene-based analyses in MAGMA v1.08^7^. Specifically, we conducted the gene-based test, which simply assigns SNPs to genes based on physical location. We also performed the competitive gene-set analysis, which uses curated gene sets and GO terms from MsigDB^8^. Finally, we conducted gene-property analyses in MAGMA, which test the relationship between tissue-specific gene expression profiles and disease-gene associations. The gene-property analyses are performed using the average expression of genes per tissue type as a gene covariate, where the gene expression values are log2 transformed average RPKM per tissue type after winsorization based on GTEx RNA-seq data. This test was performed in FUMA using the result of the MAGMA gene analysis (i.e., the gene-based p-values) and a one-sided test (testing for greater association between disease-gene associations and tissue specificity), conditioning on average expression across all tissue types.

***Differential Gene Expression***

For differential gene expression analyses, we used whole-genome transcriptomic data from Kapoor et al.^9^, specifically comparing differential gene expression in the prefrontal cortex (PFC) of 65 alcoholics versus 73 controls (post-mortem brain samples from New South Wales Tissue Resource Centre at the University of Sydney (http://sydney.edu.au/medicine/pathology/btrc/)) and in the PFC of 258 SCZ cases versus 279 controls (data obtained from the CommonMind consortium.) We used Fisher’s exact test to determine evidence of enrichment^10^.

***GeNetic cOVariance Analyzer (GNOVA)***

GNOVA^11^ uses the method of moments to provide annotation-stratified covariance

estimates that are robust to LD and sample overlap. The MAF quartiles for annotation were determined using the 1000 Genomes Phase 3 European reference panel after filtering SNPs with MAF < 0.05. GenoCanyon^12^ is a functional annotation approach which utilizes unsupervised statistical learning to integrate genomic conservation measures and biochemical annotation data to predict functional potential across the entire genome. GenoSkyline and GenoSkyline-Plus^13,14^ utilize transcriptomic and epigenomic data from ENCODE^15^ and the Roadmap Epigenomics Project^16^ to predict tissue-specific functional regions for seven broad tissue categories: brain, cardiovascular, epithelium, gastrointestinal, immune, muscle, and other tissues; these annotations were used for the tissue-specific genetic covariance partitioning. We first filtered the summary statistics to only include common HapMap3 SNPs as described previously, removed the MHC region (chr6:26000885 - chr6:33999991), then used GNOVA to estimate the $\rho_{g}$ attributable to each category in the MAF, functionality, and tissue-specific functionality annotations.

We also included alternative annotations that were defined using GTEx^17^ data on 13 specific brain regions (created using annotation files from the LDSC cell-type specific analyses^18^: <https://github.com/bulik/ldsc/wiki/Cell-type-specific-analyses>).

***Genomic Psychiatry Cohort (GPC) Investigators***

Michele T Pato MD ^1,2^, Carlos N Pato, MD, PhD ^1,2^, Tim B Bigdeli, PhD ^1,2,3^, Ayman H Fanous, MD ^1,2,3^, Steven A McCarroll, PhD ^4,5^, Peter F Buckley, MD ^6^, Mark J. Daly ^7,8,9,5^, James A Knowles MD, PhD ^2,10^, Douglas S Lehrer, MD ^11^, Dolores Malaspina, MD, MSPH ^12,13^, Mark H Rapaport, MD ^14^, Jeffrey J Rakofsky, MD ^14^, Janet L Sobell, PhD ^15^, Giulio Genovese, PhD ^4,5^, Penelope Georgakopoulos, DrPH ^2^, Jacquelyn L Meyers, PhD ^1^, Roseann E Peterson, PhD ^6^, Helena Medeiros, MSW ^2^, Jorge Valderrama, PhD ^1,2^, Eric D Achtyes, MD ^16^, Roman Kotov, PhD ^17^, Colony Abbott, MPH ^16^, Maria Helena Azevedo, PhD ^18^, Richard A Belliveau, Jr, BA ^4^, Elizabeth Bevilacqua, BS ^19^, Evelyn J Bromet, PhD ^17^, William Byerley, MD ^20^, Celia Barreto Carvalho, PhD ^21^, Sinéad B Chapman, MS ^4^, Lynn E DeLisi, MD ^22,23^, Ashley L Dumont, BASc ^4^, Colm O'Dushlaine, PhD ^4^, Oleg V Evgrafov, PhD ^2,10^, Laura J Fochtmann, MD ^17^, Diane Gage ^4^, James L Kennedy, MD ^24^, Becky Kinkead, PhD ^14^, Antonio Macedo, PhD ^18^, Jennifer L Moran, PhD ^4^, Christopher P Morley, PhD ^25-27^, Mantosh J Dewan, MD ^27^, James Nemesh ^4^, Diana O Perkins, MD, MPH ^28^, Shaun M Purcell, PhD ^4,29^, Edward M Scolnick, MD ^4^,  Brooke M Sklar, MA ^15^, Pamela Sklar, MD, PhD ^12,13^, Jordan W Smoller, MD, ScD ^4,23,30,31^, Patrick F Sullivan, MD, FRANZCP ^28,32^, Humberto Nicolini, MD ^33^, Conrad O Iyegbe, PhD ^34^, Fabio Macciardi, MD, PhD ^35^, Stephen R Marder, MD ^36,37^, Michael A Escamilla, MD ^38^, Ruben C Gur, PhD ^39-41^, Raquel E Gur, MD, PhD ^39-41^, Tiffany A Greenwood, PhD ^42^, David L Braff, MD ^42,43^, Marquis P Vawter, PhD, MA, MS ^35^

1 Department of Psychiatry and Behavioral Sciences and ^2^ Institute for Genomic Health, SUNY Downstate Medical Center, Brooklyn, NY, USA; ^3^ VA New York Harbor Healthcare System, Brooklyn, NY, USA; ^4^ Stanley Center for Psychiatric Research, Broad Institute of MIT and Harvard, Cambridge, MA, USA; ^5^ Department of Genetics, Harvard Medical School, Boston, MA, USA; ^6^ School of Medicine, Virginia Commonwealth University, Richmond, VA, USA; ^7^ Institute for Molecular Medicine Finland (FIMM), University of Helsinki, Helsinki, Finland; ^8^ Analytic and Translational Genetics Unit, Massachusetts General Hospital, Boston, MA, USA; ^9^ Program in Medical and Population Genetics, Broad Institute of Harvard and MIT, Cambridge, MA, USA; ^10^ Department of Cell Biology, SUNY Downstate Medical Center, Brooklyn, NY, USA; ^11^ Department of Psychiatry, Wright State University, Dayton, OH, USA; ^12^ Departments of Psychiatry and ^13^ Genetics & Genomics, Icahn School of Medicine at Mount Sinai, NY, USA; ^14^ Department of Psychiatry and Behavioral Sciences, Emory University, Atlanta, GA, USA; ^15^ Department of Psychiatry & Behavioral Sciences, University of Southern California, Los Angeles, CA, USA; ^16^ Cherry Health and Michigan State University College of Human Medicine, Grand Rapids, MI, USA; ^17^ Department of Psychiatry, Stony Brook University, Stony Brook, NY, USA; ^18^ Institute of Medical Psychology, Faculty of Medicine, University of Coimbra, Coimbra, PT; ^19^ Beacon Health Options, Boston, MA, USA; ^20^ Department of Psychiatry, University of California, San Francisco, CA, USA; ^2^1 Faculty of Social and Human Sciences, University of Azores, PT; ^22^ VA Boston Healthcare System, Brockton, MA, USA; ^23^ Department of Psychiatry, Harvard Medical School, Boston, MA, USA; ^24^ Neurogenetics Laboratory, Campbell Family Mental Health Research Institute, Centre for Addiction and Mental Health; Department of Psychiatry, University of Toronto, ON, CA; ^25^ Departments of Public Health and Preventive Medicine, ^26^ Family Medicine, and ^27^ Psychiatry and Behavioral Sciences, State University of New York, Upstate Medical University, Syracuse, NY, USA; ^28^ Department of Psychiatry, University of North Carolina, Chapel Hill, NC, USA; ^29^ Department of Psychiatry, Brigham and Women’s Hospital, Boston, MA, USA; ^30^ Department of Psychiatry, Massachusetts General Hospital, Boston, MA, USA; ^31^ Department of Epidemiology, Harvard T.H. Chan School of Public Health, Boston, MA, USA; ^32^ Medical Epidemiology and Biostatistics, Karolinska Institutet, Solna, SE; ^33^ Carracci Medical Group, Mexico City, MX; ^34^ Department of Psychosis Studies, King’s College London, London, UK; ^35^ Department of Psychiatry and Human Behavior, University of California, Irvine, CA, USA; ^36^ Department of Psychiatry and Biobehavioral Sciences and ^37^ Semel Institute for Neuroscience and Human Behavior, Geffen School of Medicine, University of California Los Angeles, Los Angeles, CA, USA; ^38^ Department of Psychiatry, University of Texas Rio Grande Valley School of Medicine; ^39^ Departments of Psychiatry and ^40^ Child & Adolescent Psychiatry and ^41^ Lifespan Brain Institute, University of Pennsylvania Perelman School of Medicine and Children's Hospital of Philadelphia, Philadelphia, PA, USA;  ^42^ Department of Psychiatry, University of California, La Jolla, San Diego, CA, USA; ^43^ VISN-22 Mental Illness, Research, Education and Clinical Center (MIRECC), VA San Diego Healthcare System, San Diego, CA, USA

***MVP Core Acknowledgements***

**MVP Executive Committee**

- Co-Chair: J. Michael Gaziano, M.D., M.P.H.

VA Boston Healthcare System, 150 S. Huntington Avenue, Boston, MA 02130

- Co-Chair: Sumitra Muralidhar, Ph.D.

US Department of Veterans Affairs, 810 Vermont Avenue NW, Washington, DC 20420

- Rachel Ramoni, D.M.D., Sc.D., Chief VA Research and Development Officer

US Department of Veterans Affairs, 810 Vermont Avenue NW, Washington, DC 20420

- Jean Beckham, Ph.D.

Durham VA Medical Center, 508 Fulton Street, Durham, NC 27705

- Kyong-Mi Chang, M.D.

Philadelphia VA Medical Center, 3900 Woodland Avenue, Philadelphia, PA 19104

- Christopher J. O’Donnell, M.D., M.P.H.

VA Boston Healthcare System, 150 S. Huntington Avenue, Boston, MA 02130

- Philip S. Tsao, Ph.D.

VA Palo Alto Health Care System, 3801 Miranda Avenue, Palo Alto, CA 94304

- James Breeling, M.D., Ex-Officio

US Department of Veterans Affairs, 810 Vermont Avenue NW, Washington, DC 20420

- Grant Huang, Ph.D., Ex-Officio

US Department of Veterans Affairs, 810 Vermont Avenue NW, Washington, DC 20420

- JP Casas Romero, M.D., Ph.D., Ex-Officio

VA Boston Healthcare System, 150 S. Huntington Avenue, Boston, MA 02130

**MVP Program Office**

- Sumitra Muralidhar, Ph.D.

US Department of Veterans Affairs, 810 Vermont Avenue NW, Washington, DC 20420

- Jennifer Moser, Ph.D.

US Department of Veterans Affairs, 810 Vermont Avenue NW, Washington, DC 20420

**MVP Recruitment/Enrollment**

- Recruitment/Enrollment Director/Deputy Director, Boston – Stacey B. Whitbourne, Ph.D.; Jessica V. Brewer, M.P.H.

VA Boston Healthcare System, 150 S. Huntington Avenue, Boston, MA 02130

- MVP Coordinating Centers
  - Clinical Epidemiology Research Center (CERC), West Haven – Mihaela Aslan, Ph.D.

West Haven VA Medical Center, 950 Campbell Avenue, West Haven, CT 06516

- Cooperative Studies Program Clinical Research Pharmacy Coordinating Center, Albuquerque – Todd Connor, Pharm.D.; Dean P. Argyres, B.S., M.S.

New Mexico VA Health Care System, 1501 San Pedro Drive SE, Albuquerque, NM 87108

- Genomics Coordinating Center, Palo Alto – Philip S. Tsao, Ph.D.

VA Palo Alto Health Care System, 3801 Miranda Avenue, Palo Alto, CA 94304

- MVP Boston Coordinating Center, Boston - J. Michael Gaziano, M.D., M.P.H.

VA Boston Healthcare System, 150 S. Huntington Avenue, Boston, MA 02130

- MVP Information Center, Canandaigua – Brady Stephens, M.S.

Canandaigua VA Medical Center, 400 Fort Hill Avenue, Canandaigua, NY 14424

- VA Central Biorepository, Boston – Mary T. Brophy M.D., M.P.H.; Donald E. Humphries, Ph.D.; Luis E. Selva, Ph.D.

VA Boston Healthcare System, 150 S. Huntington Avenue, Boston, MA 02130

- MVP Informatics, Boston – Nhan Do, M.D.; Shahpoor Shayan

VA Boston Healthcare System, 150 S. Huntington Avenue, Boston, MA 02130

- MVP Data Operations/Analytics, Boston – Kelly Cho, Ph.D.

VA Boston Healthcare System, 150 S. Huntington Avenue, Boston, MA 02130

**MVP Science**

- Science Operations – Christopher J. O’Donnell, M.D., M.P.H.

VA Boston Healthcare System, 150 S. Huntington Avenue, Boston, MA 02130

- Genomics Core - Christopher J. O’Donnell, M.D., M.P.H.; Saiju Pyarajan Ph.D.

VA Boston Healthcare System, 150 S. Huntington Avenue, Boston, MA 02130

Philip S. Tsao, Ph.D.

VA Palo Alto Health Care System, 3801 Miranda Avenue, Palo Alto, CA 94304

- Phenomics Core- Kelly Cho, M.P.H, Ph.D.

VA Boston Healthcare System, 150 S. Huntington Avenue, Boston, MA 02130

- Data and Computational Sciences – Saiju Pyarajan, Ph.D.

VA Boston Healthcare System, 150 S. Huntington Avenue, Boston, MA 02130

- Statistical Genetics – Elizabeth Hauser, Ph.D.

Durham VA Medical Center, 508 Fulton Street, Durham, NC 27705

Yan Sun, Ph.D.

Atlanta VA Medical Center, 1670 Clairmont Road, Decatur, GA 30033

Hongyu Zhao, Ph.D.

West Haven VA Medical Center, 950 Campbell Avenue, West Haven, CT 06516

**Current MVP Local Site Investigators**

- Atlanta VA Medical Center (Peter Wilson, M.D.)

1670 Clairmont Road, Decatur, GA 30033

- Bay Pines VA Healthcare System (Rachel McArdle, Ph.D.)

10,000 Bay Pines Blvd Bay Pines, FL 33744

- Birmingham VA Medical Center (Louis Dellitalia, M.D.)

700 S. 19th Street, Birmingham AL 35233

- Central Western Massachusetts Healthcare System (Kristin Mattocks, Ph.D., M.P.H.)

421 North Main Street, Leeds, MA 01053

- Cincinnati VA Medical Center (John Harley, M.D., Ph.D.)

3200 Vine Street, Cincinnati, OH 45220

- Clement J. Zablocki VA Medical Center (Jeffrey Whittle, M.D., M.P.H.)

5000 West National Avenue, Milwaukee, WI 53295

- VA Northeast Ohio Healthcare System (Frank Jacono, M.D.)

10701 East Boulevard, Cleveland, OH 44106

- Durham VA Medical Center (Jean Beckham, Ph.D.)

508 Fulton Street, Durham, NC 27705

- Edith Nourse Rogers Memorial Veterans Hospital (John Wells., Ph.D.)

200 Springs Road, Bedford, MA 01730

- Edward Hines, Jr. VA Medical Center (Salvador Gutierrez, M.D.)

5000 South 5th Avenue, Hines, IL 60141

- Veterans Health Care System of the Ozarks (Gretchen Gibson, D.D.S., M.P.H.)

1100 North College Avenue, Fayetteville, AR 72703

- Fargo VA Health Care System (Kimberly Hammer, Ph.D.)

2101 N. Elm, Fargo, ND 58102

- VA Health Care Upstate New York (Laurence Kaminsky, Ph.D.)

113 Holland Avenue, Albany, NY 12208

- New Mexico VA Health Care System (Gerardo Villareal, M.D.)

1501 San Pedro Drive, S.E. Albuquerque, NM 87108

- VA Boston Healthcare System (Scott Kinlay, M.B.B.S., Ph.D.)

150 S. Huntington Avenue, Boston, MA 02130

- VA Western New York Healthcare System (Junzhe Xu, M.D.)

3495 Bailey Avenue, Buffalo, NY 14215-1199

- Ralph H. Johnson VA Medical Center (Mark Hamner, M.D.)

109 Bee Street, Mental Health Research, Charleston, SC 29401

- Columbia VA Health Care System (Roy Mathew, M.D.)

6439 Garners Ferry Road, Columbia, SC 29209

- VA North Texas Health Care System (Sujata Bhushan, M.D.)

4500 S. Lancaster Road, Dallas, TX 75216

- Hampton VA Medical Center (Pran Iruvanti, D.O., Ph.D.)

100 Emancipation Drive, Hampton, VA 23667

- Richmond VA Medical Center (Michael Godschalk, M.D.)

1201 Broad Rock Blvd., Richmond, VA 23249

- Iowa City VA Health Care System (Zuhair Ballas, M.D.)

601 Highway 6 West, Iowa City, IA 52246-2208

- Eastern Oklahoma VA Health Care System (Douglas Ivins, M.D.)

1011 Honor Heights Drive, Muskogee, OK 74401

- James A. Haley Veterans’ Hospital (Stephen Mastorides, M.D.)

13000 Bruce B. Downs Blvd, Tampa, FL 33612

- James H. Quillen VA Medical Center (Jonathan Moorman, M.D., Ph.D.)

Corner of Lamont & Veterans Way, Mountain Home, TN 37684

- John D. Dingell VA Medical Center (Saib Gappy, M.D.)

4646 John R Street, Detroit, MI 48201

- Louisville VA Medical Center (Jon Klein, M.D., Ph.D.)

800 Zorn Avenue, Louisville, KY 40206

- Manchester VA Medical Center (Nora Ratcliffe, M.D.)

718 Smyth Road, Manchester, NH 03104

- Miami VA Health Care System (Hermes Florez, M.D., Ph.D.)

1201 NW 16th Street, 11 GRC, Miami FL 33125

- Michael E. DeBakey VA Medical Center (Olaoluwa Okusaga, M.D.)

2002 Holcombe Blvd, Houston, TX 77030

- Minneapolis VA Health Care System (Maureen Murdoch, M.D., M.P.H.)

One Veterans Drive, Minneapolis, MN 55417

- N. FL/S. GA Veterans Health System (Peruvemba Sriram, M.D.)

1601 SW Archer Road, Gainesville, FL 32608

- Northport VA Medical Center (Shing Shing Yeh, Ph.D., M.D.)

79 Middleville Road, Northport, NY 11768

- Overton Brooks VA Medical Center (Neeraj Tandon, M.D.)

510 East Stoner Ave, Shreveport, LA 71101

- Philadelphia VA Medical Center (Darshana Jhala, M.D.)

3900 Woodland Avenue, Philadelphia, PA 19104

- Phoenix VA Health Care System (Samuel Aguayo, M.D.)

650 E. Indian School Road, Phoenix, AZ 85012

- Portland VA Medical Center (David Cohen, M.D.)

3710 SW U.S. Veterans Hospital Road, Portland, OR 97239

- Providence VA Medical Center (Satish Sharma, M.D.)

830 Chalkstone Avenue, Providence, RI 02908

- Richard Roudebush VA Medical Center (Suthat Liangpunsakul, M.D., M.P.H.)

1481 West 10th Street, Indianapolis, IN 46202

- Salem VA Medical Center (Kris Ann Oursler, M.D.)

1970 Roanoke Blvd, Salem, VA 24153

- San Francisco VA Health Care System (Mary Whooley, M.D.)

4150 Clement Street, San Francisco, CA 94121

- South Texas Veterans Health Care System (Sunil Ahuja, M.D.)

7400 Merton Minter Boulevard, San Antonio, TX 78229

- Southeast Louisiana Veterans Health Care System (Joseph Constans, Ph.D.)

2400 Canal Street, New Orleans, LA 70119

- Southern Arizona VA Health Care System (Paul Meyer, M.D., Ph.D.)

3601 S 6th Avenue, Tucson, AZ 85723

- Sioux Falls VA Health Care System (Jennifer Greco, M.D.)

2501 W 22nd Street, Sioux Falls, SD 57105

- St. Louis VA Health Care System (Michael Rauchman, M.D.)

915 North Grand Blvd, St. Louis, MO 63106

- Syracuse VA Medical Center (Richard Servatius, Ph.D.)

800 Irving Avenue, Syracuse, NY 13210

- VA Eastern Kansas Health Care System (Melinda Gaddy, Ph.D.)

4101 S 4th Street Trafficway, Leavenworth, KS 66048

- VA Greater Los Angeles Health Care System (Agnes Wallbom, M.D., M.S.)

11301 Wilshire Blvd, Los Angeles, CA 90073

- VA Long Beach Healthcare System (Timothy Morgan, M.D.)

5901 East 7th Street Long Beach, CA 90822

- VA Maine Healthcare System (Todd Stapley, D.O.)

1 VA Center, Augusta, ME 04330

- VA New York Harbor Healthcare System (Scott Sherman, M.D., M.P.H.)

423 East 23rd Street, New York, NY 10010

- VA Pacific Islands Health Care System (George Ross, M.D.)

459 Patterson Rd, Honolulu, HI 96819

- VA Palo Alto Health Care System (Philip Tsao, Ph.D.)

3801 Miranda Avenue, Palo Alto, CA 94304-1290

- VA Pittsburgh Health Care System (Patrick Strollo, Jr., M.D.)

University Drive, Pittsburgh, PA 15240

- VA Puget Sound Health Care System (Edward Boyko, M.D.)

1660 S. Columbian Way, Seattle, WA 98108-1597

- VA Salt Lake City Health Care System (Laurence Meyer, M.D., Ph.D.)

500 Foothill Drive, Salt Lake City, UT 84148

- VA San Diego Healthcare System (Samir Gupta, M.D., M.S.C.S.)

3350 La Jolla Village Drive, San Diego, CA 92161

- VA Sierra Nevada Health Care System (Mostaqul Huq, Pharm.D., Ph.D.)

975 Kirman Avenue, Reno, NV 89502

- VA Southern Nevada Healthcare System (Joseph Fayad, M.D.)

6900 North Pecos Road, North Las Vegas, NV 89086

- VA Tennessee Valley Healthcare System (Adriana Hung, M.D., M.P.H.)

1310 24th Avenue, South Nashville, TN 37212

- Washington DC VA Medical Center (Jack Lichy, M.D., Ph.D.)

50 Irving St, Washington, D. C. 20422

- W.G. (Bill) Hefner VA Medical Center (Robin Hurley, M.D.)

1601 Brenner Ave, Salisbury, NC 28144

- White River Junction VA Medical Center (Brooks Robey, M.D.)

163 Veterans Drive, White River Junction, VT 05009

- William S. Middleton Memorial Veterans Hospital (Robert Striker, M.D., Ph.D.)

2500 Overlook Terrace, Madison, WI 53705
