## Supplemental Figures for "Investigation of convergent and divergent genetic influences underlying schizophrenia and alcohol use disorder"

**Fig S1. Conceptual overview of ASSET analysis.**

**Fig S2. MAGMA gene-based Manhattan plots of ASSET output for European -ancestry cross-disorder.**

**Fig S3. MAGMA gene-property tissue expression results for pleiotropic variants in the European-ancestry cross-disorder.**

**Fig S4. Results of the brain region-specific LDSC-SEG analyses.**

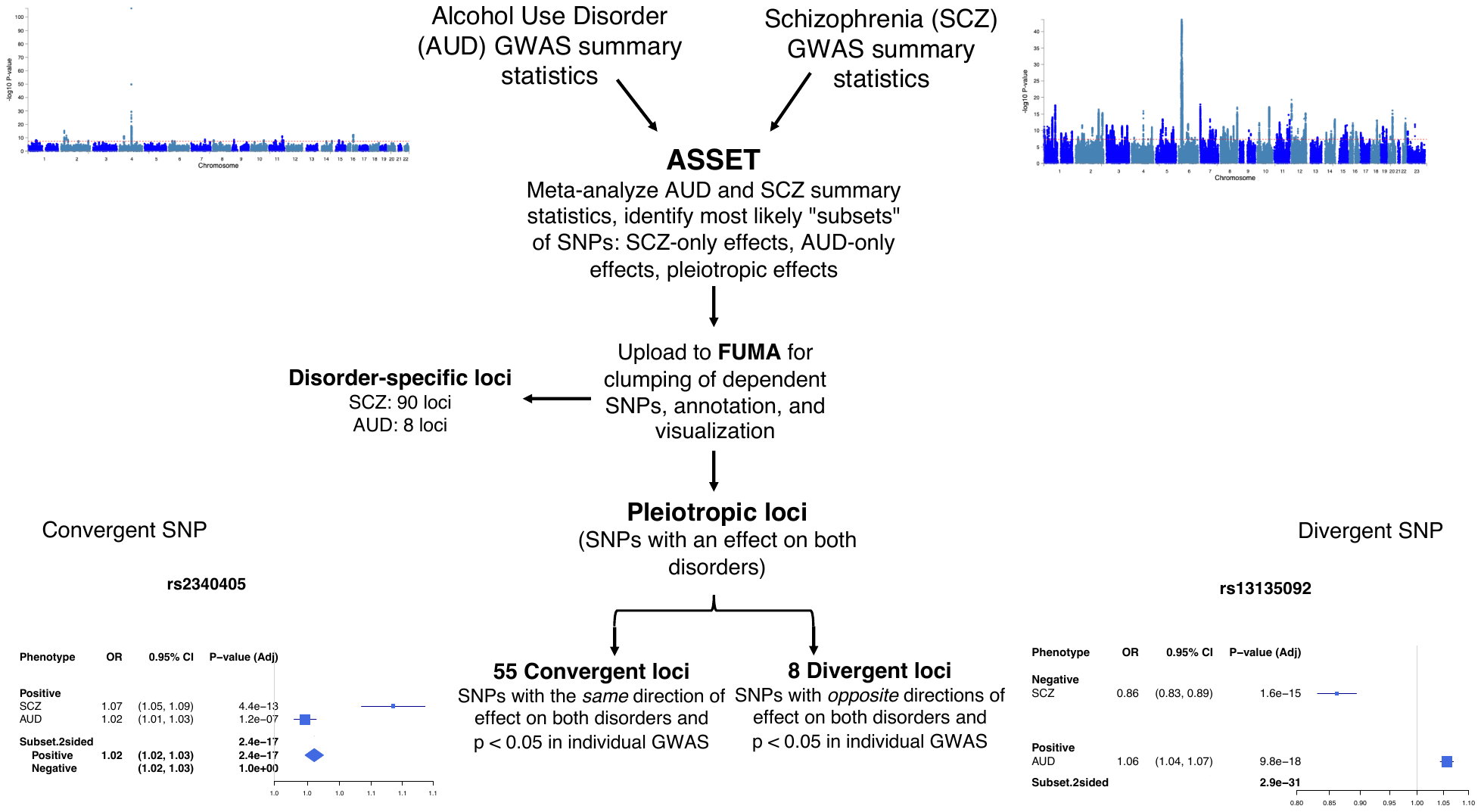

**Fig S1. Conceptual overview of ASSET analysis of AUD and SCZ**

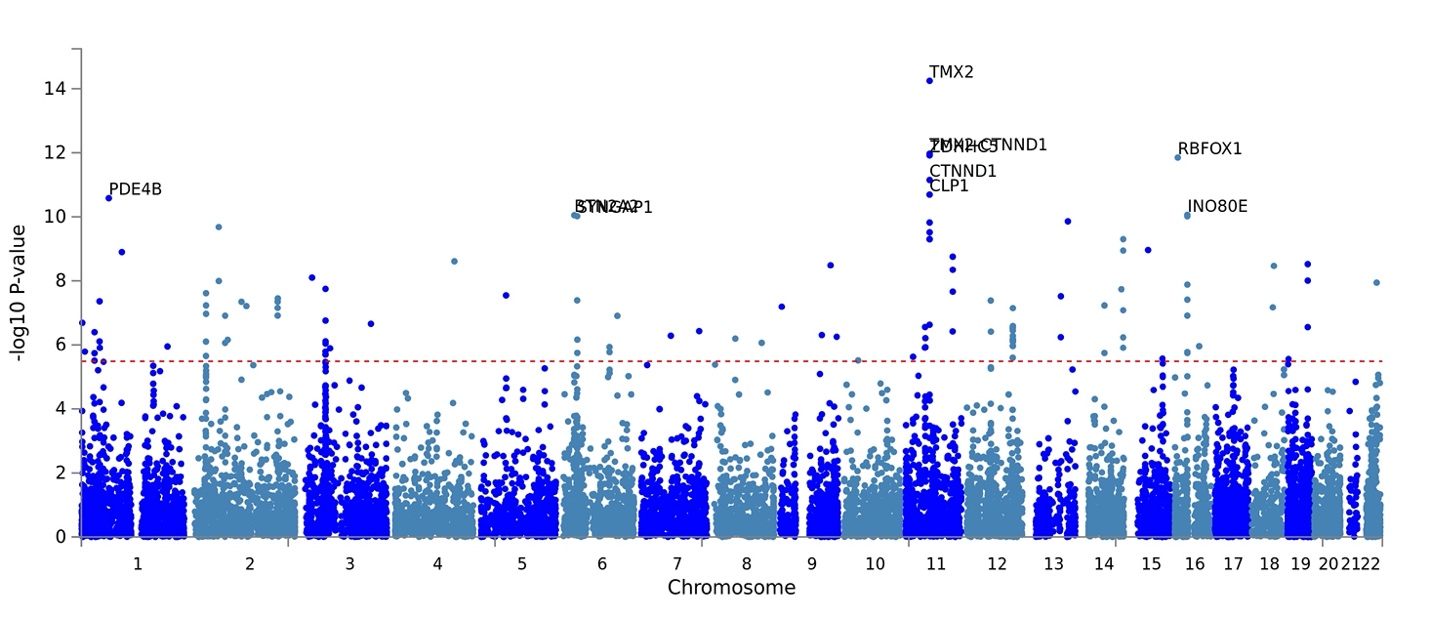

a. **MAGMA gene-based results for the convergent subset of ASSET output (variants with the same direction of effect on both AUD and SCZ).** Only the top 10 genes are shown for clarity.

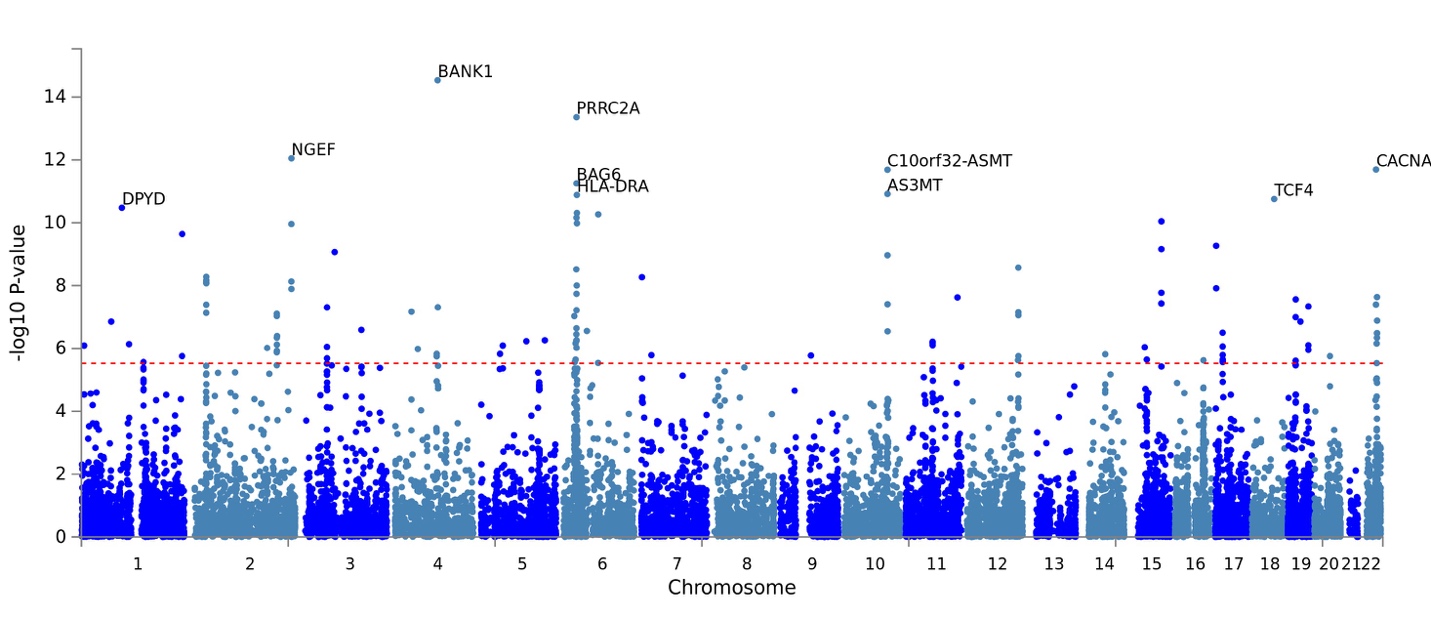

b. **MAGMA gene-based results for the divergent subset of ASSET output (variants with opposite directions of effect on AUD and SCZ).** Only the top 10 genes are shown for clarity.

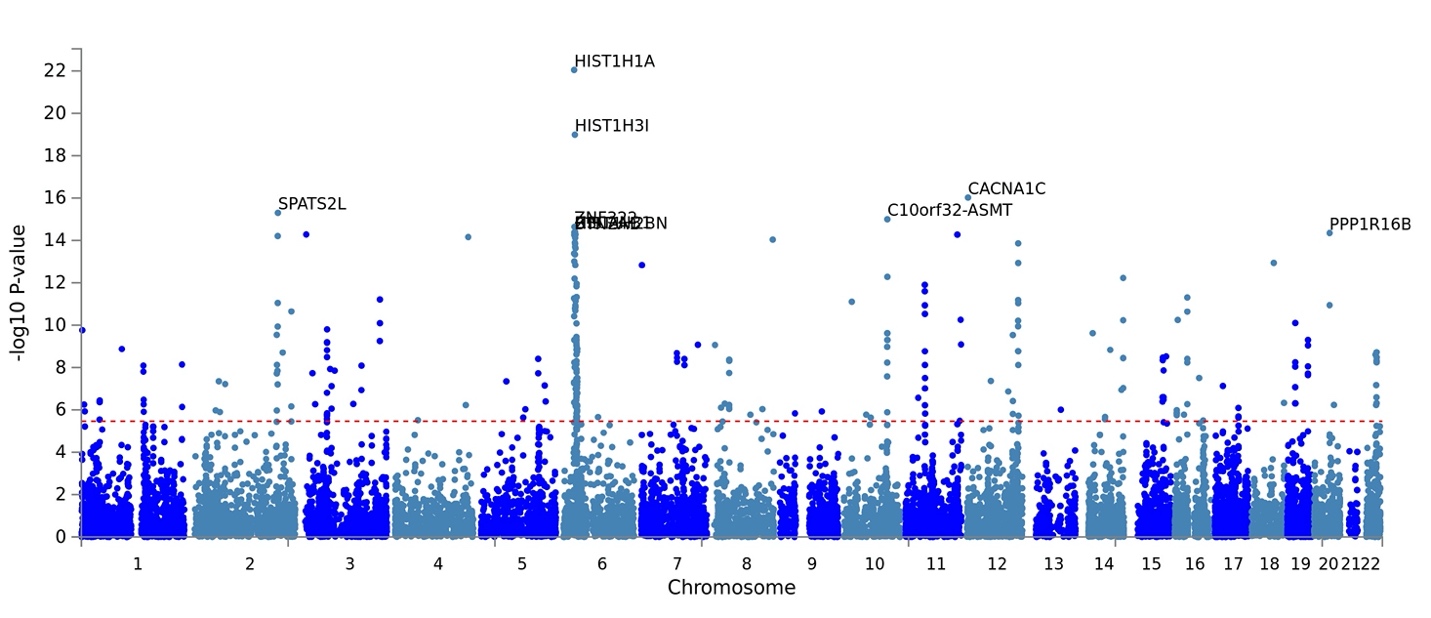

c. **MAGMA gene-based results for the SCZ-specific subset of ASSET output.** Only the top 10 genes are shown for clarity.

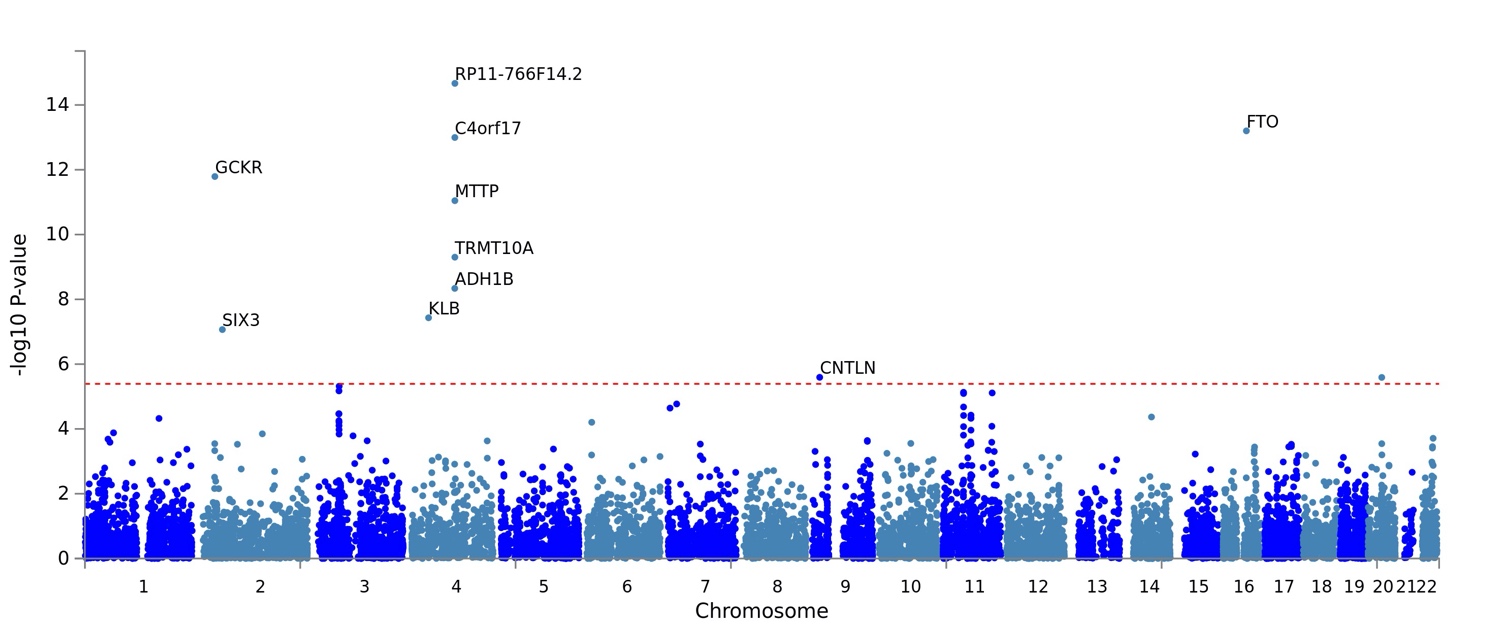

d. **MAGMA gene-based results for the AUD-specific subset of ASSET output.** Only the top 10 genes are shown for clarity.

**Figure S2. MAGMA v1.08 gene-based Manhattan plots from ASSET EUR-ancestry summary statistics.** Plots downloaded from FUMA. The dotted line indicates significance threshold, correcting for the number of genes tested. Only the top 10 genes are labeled.

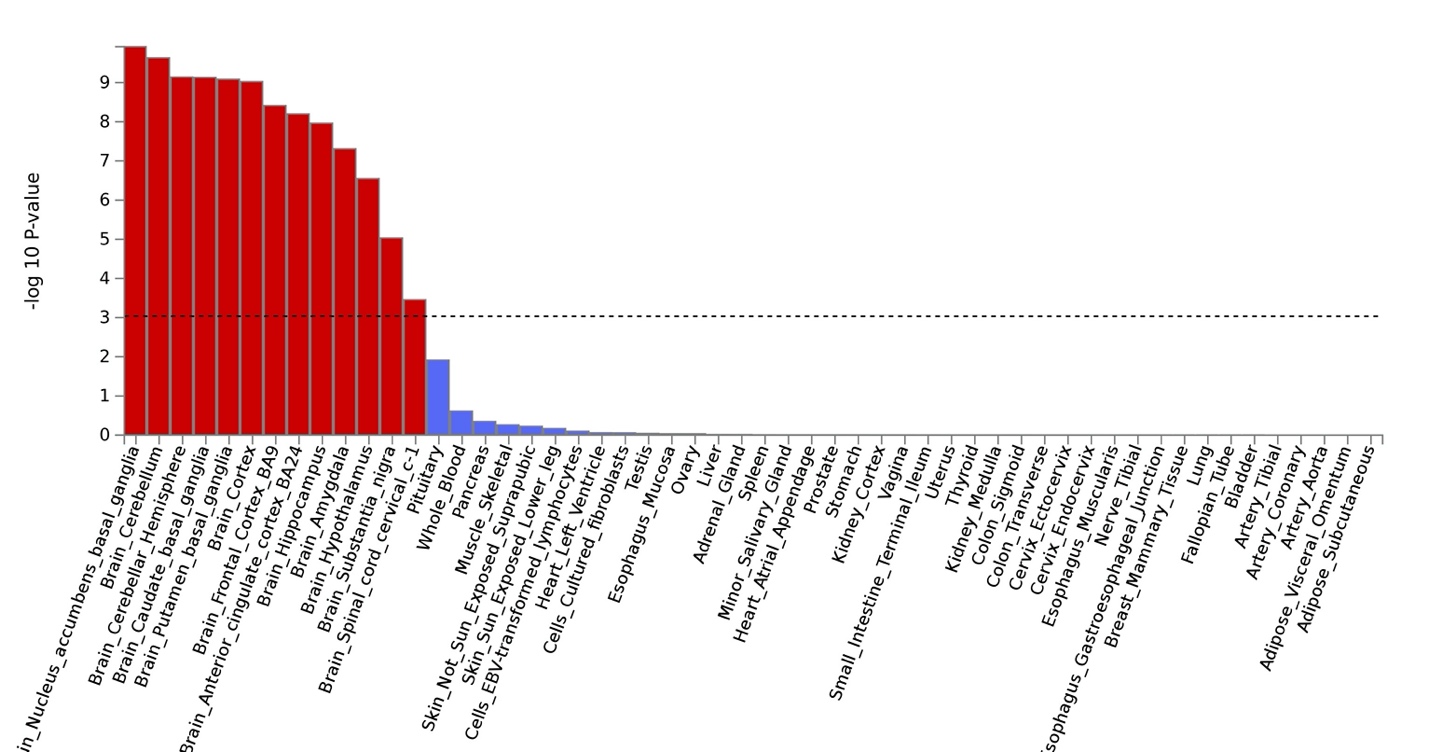

a. **MAGMA gene-property results for the convergent subset of variants based on GTEx v8 53 tissues**

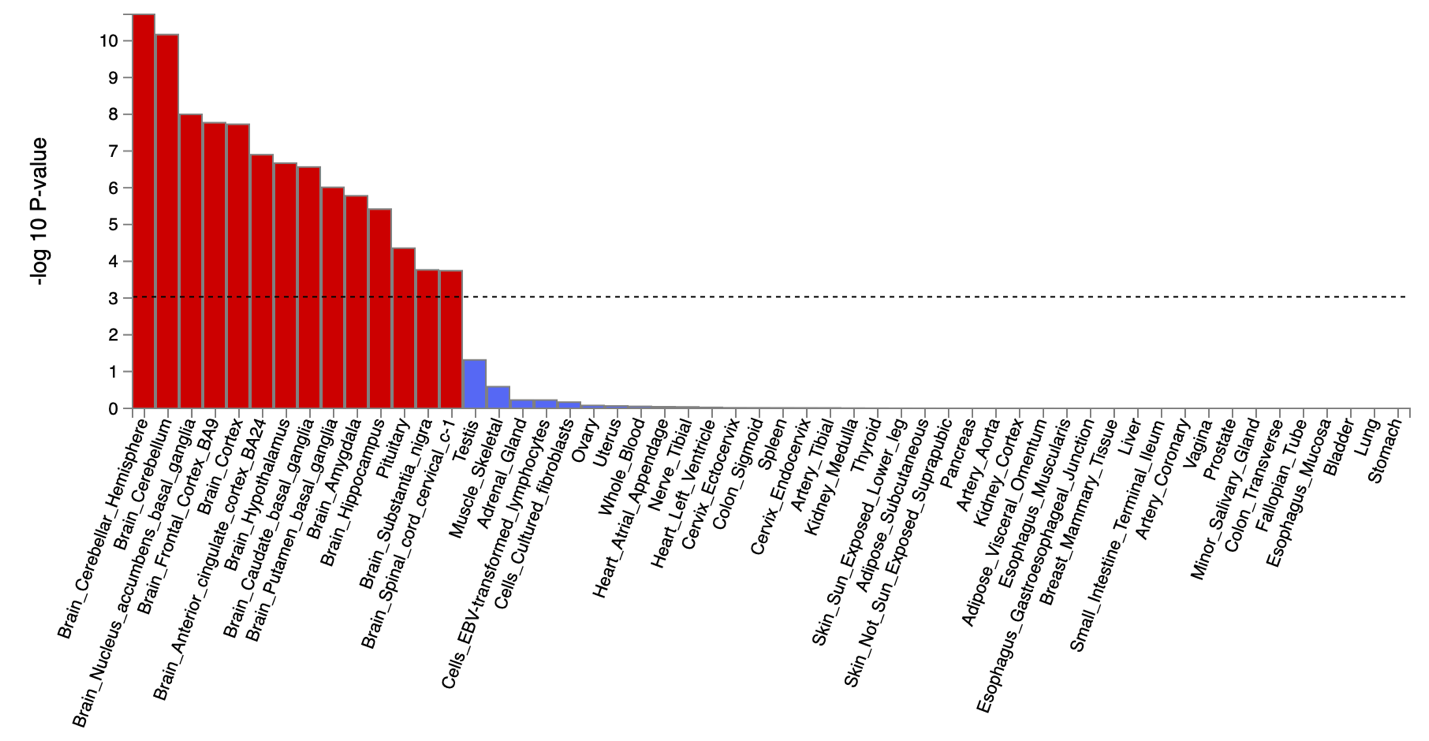

b. **MAGMA gene-property results for the divergent subset of variants based on GTEx v8 53 tissues**

**Figure S3. MAGMA v1.08 gene-property tissue and age-specific expression results for pleiotropic variants in the European-ancestry cross-disorder.** Plots downloaded from the FUMA v1.36a platform.

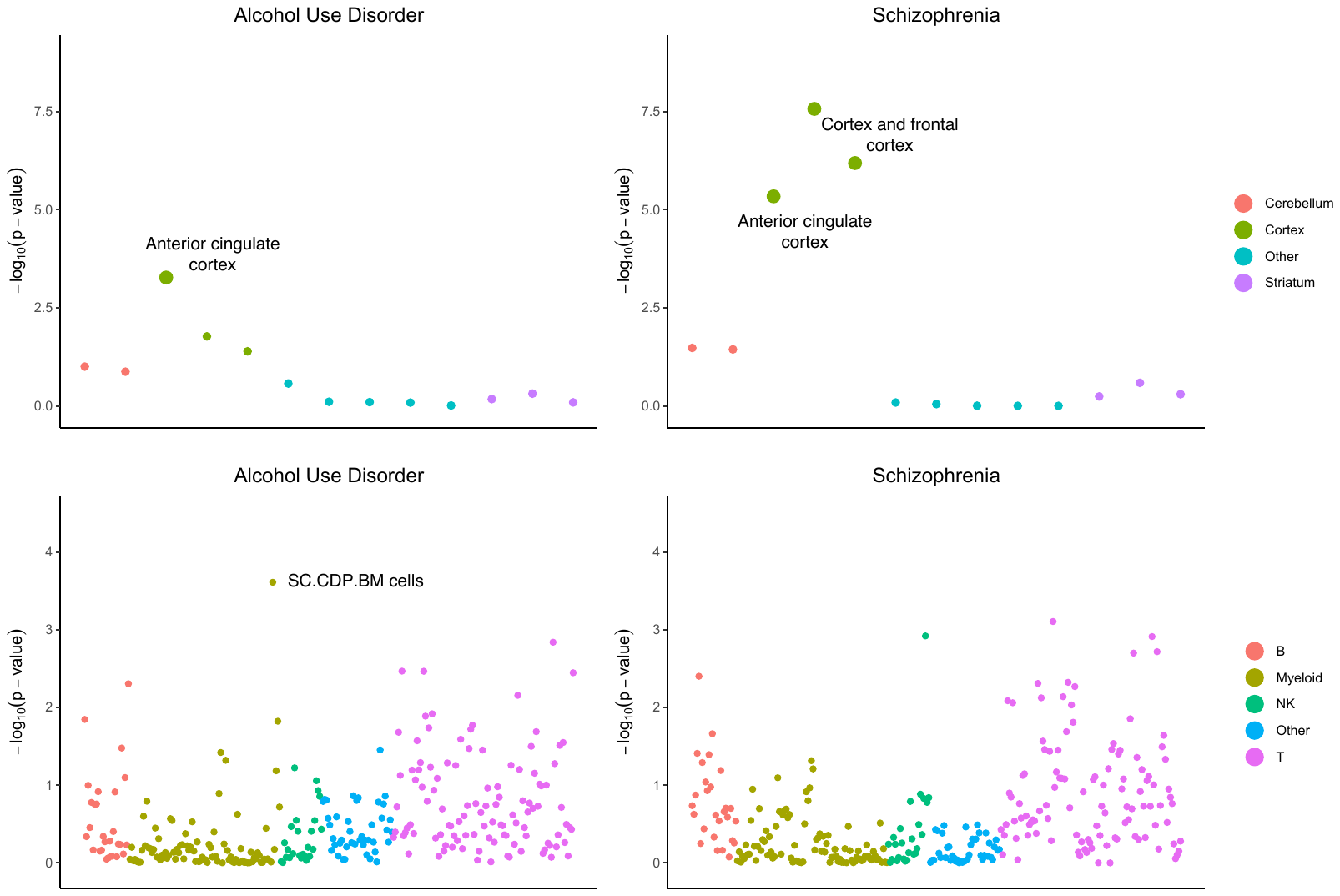

**Figure S4. Results of the brain region-specific LDSC-SEG analyses.** LDSC-SEG analysis of 13 brain regions in GTEx v6 gene expression data, classified into four groups for AUD (left) and SCZ (right). Large circles passed the cutoff of FDR < 5%.
